# Blood RNA signatures outperform CRP triage of tuberculosis lymphadenitis and pericarditis

**DOI:** 10.1101/2024.06.21.24309099

**Authors:** Tiffeney Mann, Stephanie Minnies, Rishi K Gupta, Byron WP Reeve, Georgina Nyawo, Zaida Palmer, Charissa Naidoo, Anton Doubell, Alfonso Pecararo, Thadathilankal-Jess John, Pawel Schubert, Claire J Calderwood, Aneesh Chandran, Grant Theron, Mahdad Noursadeghi

## Abstract

**Background:** Limited data are available on the diagnostic accuracy of blood RNA biomarker signatures for extrapulmonary TB (EPTB). We addressed this question among people investigated for TB lymphadenitis and TB pericarditis, in Cape Town, South Africa.

**Methods:** We enrolled 440 consecutive adults referred to a hospital for invasive sampling for presumptive TB lymphadenitis (n=300) or presumptive TB pericarditis (n=140). Samples from the site of disease underwent culture and/or molecular testing for *Mycobacterium tuberculosis* complex (Mtb). Discrimination of patients with and without TB defined by microbiology or cytology reference standards was evaluated using seven previously reported blood RNA signatures by area under the receiver-operating characteristic curve (AUROC) and sensitivity/specificity at predefined thresholds, benchmarked against blood C-reactive protein (CRP) and the World Health Organization (WHO) target product profile (TPP) for a TB triage test. Decision curve analysis (DCA) was used to evaluate the clinical utility of the best performing blood RNA signature and CRP.

**Results:** Data from 374 patients for whom results were available from at least one microbiological test from the site of disease, and blood CRP and RNA measurements, were included. Using microbiological results as the reference standard in the primary analysis (N=204 with TB), performance was similar across lymphadenitis and pericarditis patients. In the pooled analysis of both cohorts, all RNA signatures had comparable discrimination with AUROC point estimates ranging 0.77-0.82, superior to that of CRP (0.61, 95% confidence interval 0.56-0.67). The best performing signature (Roe3) achieved an AUROC of 0.82 (0.77-0.86). At a predefined threshold of 2 standard deviations (Z2) above the mean of a healthy reference control group, this signature achieved 78% (72-83%) sensitivity and 69% (62-75%) specificity. In this setting, DCA revealed that Roe3 offered greater net benefit than other approaches for services aiming to reduce the number needed to investigate with confirmatory testing to <4 to identify each case of TB.

**Interpretation:** RNA biomarkers show better accuracy and clinical utility than CRP to trigger confirmatory TB testing in patients with TB lymphadenitis and TB pericarditis, but still fall short of the WHO TPP for TB triage tests.

**Funding:** South African MRC, EDCTP2, NIH/NIAID, Wellcome Trust, NIHR, Royal College of Physicians London.

**Research in context:** *Evidence before this study:* Blood RNA biomarker signatures and CRP measurements have emerged as potential triage tests for TB, but evidence is mostly limited to their performance in pulmonary TB. Microbiological diagnosis of extrapulmonary TB (EPTB) is made challenging by the need for invasive sampling to obtain tissue from the site of disease. This is compounded by lower sensitivity of confirmatory molecular tests for EPTB compared to their performance in pulmonary disease. We performed a systematic review of diagnostic accuracy studies of blood RNA biomarkers or CRP measurements for EPTB, which could mitigate the need for site-of-disease sampling for the diagnosis of TB. We searched PubMed up to 1^st^ August 2023, using the following criteria: “extrapulmonary [title/abstract] AND tuberculosis [title/abstract] AND biomarker [title/abstract]”. Although extrapulmonary TB was included in several studies, none focused specifically on EPTB or included an adequate number of EPTB cases to provide precise estimates of test accuracy.

*Added value of this study:* To the best of our knowledge, we report the first diagnostic accuracy study of blood RNA biomarkers and CRP for TB among people with EPTB syndromes. We examined the performance of seven previously identified blood RNA biomarkers as triage tests for TB lymphadenitis and TB pericarditis compared to a microbiology reference standard among people referred to hospital for invasive sampling in a high TB and HIV prevalence setting. Multiple blood RNA biomarkers showed comparable diagnostic accuracy to that previously reported for pulmonary TB in both EPTB disease cohorts, irrespective of HIV status. All seven blood RNA biomarkers showed superior diagnostic accuracy to CRP for both lymphadenitis and pericarditis, but failed to meet the combined >90% sensitivity and >70% specificity recommended for a blood-based diagnostic triage test by WHO. Nonetheless, in decision curve analysis, an approach of using the best performing blood RNA biomarker to trigger confirmatory microbiological testing showed superior clinical utility in clinical services seeking to reduce the number needed to test (using invasive confirmatory testing) to less than 4 for each EPTB case detected. If acceptable to undertake invasive testing in more than 4 people for each true case detected, then a test-all approach will provide greater net benefit in this TB/HIV hyperendemic setting.

*Implications of all the available evidence:* Blood RNA biomarkers show some potential as diagnostic triage tests for TB lymphadenitis and TB pericarditis, but do not provide the level of accuracy for blood-based triage tests recommended by WHO for community-based tests. CRP has inferior diagnostic accuracy to blood RNA biomarkers and cannot be recommended for diagnostic triage among people with EPTB syndromes referred for invasive sampling.

## Introduction

Delays in the diagnosis of tuberculosis (TB) remain a significant barrier to reducing the associated morbidity and mortality for individuals, and effectiveness of world-wide TB control programmes at population level^1^. It is estimated that extrapulmonary disease comprises approximately 10-30% of new or relapse TB cases^1,2^. Whilst TB incidence has slowly declined over the past 20 years, EPTB incidence has not^3–5^. Microbiological diagnosis of TB has mostly relied on sampling the site of disease. In pulmonary TB, non-invasive sputum expectoration can provide appropriate samples for microbiological diagnosis. Diagnosis of extrapulmonary TB (EPTB) remains more challenging because of the need for invasive procedures to sample the site of disease, necessitating access to hospitals with specialised infrastructure and expertise that are resource intensive. In addition, confirmatory molecular tests for TB have lower sensitivity in EPTB compared to pulmonary disease^6,7^. These barriers have the greatest impact in low and middle income countries (LMIC) where EPTB incidence may be systematically underestimated, particularly among people living with HIV (PLWH) who have higher risk of EPTB^8,9^. To address this limitation in both EPTB and sputum scarce pulmonary TB, and to mitigate against the health economic barriers to systematic microbiological testing, the World Health Organization (WHO) have recommended development of sensitive, cheap, point of care blood-based triage tests to enable more precise targeting of patients for confirmatory diagnostic testing^10^.

Blood RNA biomarkers and C-reactive protein (CRP) measurements have emerged as leading candidates for blood-based triage tests for TB^11,12^. CRP is a hepatocyte-derived acute phase protein. Increased levels in the circulation are induced by interleukin (IL)-6 and other acute phase cytokine activity. It is most widely used as non-specific biomarker of bacterial infections^13^. The advent of a cheap quantitative point of care test for CRP has made it a particularly attractive as a triage test for TB in LMIC settings, where it has been evaluated for screening for pulmonary TB in PLWH before starting antiretroviral therapy^14^ and among people presenting with TB symptoms^15^. The diagnostic accuracy of CRP for EPTB has not been tested.

Numerous candidate blood RNA biomarkers of TB have been reported. They range from single gene transcripts to multi-gene signatures. We have shown that the best performing blood RNA biomarkers of TB are co-correlated and reflect canonical host immune responses downstream of tumour necrosis factor (TNF) and interferon (IFN) signalling^16^. Although previous diagnostic accuracy studies of TB blood RNA biomarkers have included people with EPTB, they were not powered to evaluate differences in biomarker accuracy between pulmonary and extrapulmonary disease. No studies to date have evaluated diagnostic accuracy exclusively among people with signs and symptoms suggestive of EPTB.

EPTB can affect any organ system. On this spectrum, TB lymphadenitis is among the commonest extra-thoracic sites of disease^2–4^. TB pericarditis is less common but provides a prototypic example of the association of EPTB with HIV co-infection^17,18^. We therefore performed head-to-head comparison of the best performing candidate RNA biomarkers from our previous studies^11,16^ among people being evaluated for TB lymphadenitis or pericarditis at a referral centre in a hyper-endemic setting for TB and HIV infection. We benchmarked RNA biomarker performance against CRP and the WHO target product profile (TPP) for a blood-based triage test aiming to achieve sensitivity of ≥90% and specificity of ≥70%^10^ and evaluated comparative clinical utility by decision curve analysis.

## Methods

### Regulatory approvals and reporting standards

The study was approved by the Stellenbosch University Faculty of Health Sciences Research Ethics Committee (N16/04/050) and the Western Cape Department of Health, South Africa (WC_2016RP15_762). The study is reported in line with standards for reporting diagnostic accuracy studies (STARD) guidance^19^.

### Study cohort

Consecutive adults referred for routine investigation by invasive sampling for TB lymphadenitis (January 2017 - March 2019) or TB pericarditis (November 2016 - January 2022) were recruited from a tertiary hospital in Cape Town (South Africa). For the lymphadenitis cohort, patients who had received TB treatment within 2 months prior to presentation were excluded. For the pericarditis cohort, we included people up to 2 weeks after TB treatment initiation. Subsets of the patients included in the present study contributed to a diagnostic accuracy study of Xpert MTB/RIF Ultra (Ultra) in TB lymphadenitis^20^, and evaluation of microbiota in TB lymphadenitis^21^ or TB pericarditis^22^. Demographic characteristics, laboratory confirmed HIV status and self-reported TB treatment (within 12 weeks of enrolment) were recorded for all participants. In each cohort, diagnostic testing on samples from the site of disease included Xpert MTB/RIF (Xpert) or Ultra, with or without liquid culture for *Mycobacterium tuberculosis* (Mtb), and cytology performed on lymph node aspirates according to pre-defined algorithms (Supplementary Figure 1). For both cohorts, the results from sputum smear microscopy, Mtb culture and molecular testing on other non-site of disease fluids conducted as part of routine care were also recorded where available. Except for site-of-disease Ultra, all other investigations were performed as part of routine care.

### Measurement of blood RNA biomarkers and CRP

Serum and blood RNA (collected in Tempus™ tubes; Ambion Life Technologies) samples were taken at recruitment and stored at −80°C. Seven concise blood RNA signatures selected from the best performing biomarkers of TB in our previous comparative studies^11,16^ were measured using the Nanostring™ platform as described previously^23^. Each signature was named using the first author’s surname from the original publication where it was derived followed by the number of component genes, except for BATF2 (a single transcript) and RISK6 (named by the original investigators). These included the following signatures: BATF2^24^, Gliddon3^25^, RISK6^26^, Roe3^27^, Suliman4^28^, Sweeney3^29^ and Darboe11 (derived from Zak16)^30^. Blood RNA signature Z-scores were calculated by subtracting the mean and dividing by the standard deviation of blood RNA samples (N=48) from a multi-ethnic healthy control population of individuals with latent TB^23,31^, measured using the same Nanostring code-set batches. All samples were processed across two code-set batches. Distributions of the processed gene level data across the two code-set batches suggested a small batch effect (Supplementary Figure 2). We therefore performed batch correction with the ComBat function from the sva package in our primary analyses, as previously^23^ (Supplementary Figure 3-4). Serum CRP was measured on biobanked serum using the Cobas high-sensitive immunoturbidimetric assay (Roche Diagnostics Limited, Burgess Hill, UK).

### Reference standards

Our primary reference standard for TB in both cohorts was a positive Mtb culture or molecular test (Xpert or Ultra, excluding ‘trace’ results) on samples from the site of disease or sputum. In sensitivity analyses, we examined a range of alternative reference standards where we: (a) included Ultra ‘trace’ results as positive; (b) included patients commenced on TB treatment within 12 weeks as TB; and (c) included cytology or histology consistent with TB, as defined by evidence of necrotising granulomas.

### Primary analysis

All analyses were conducted using R (version 4.0.2). From the parent cohorts, we excluded participants without RNA (n=58), CRP (n=6) or primary reference standard (n=2) data. In our primary analysis, we quantified area under the receiver operating characteristic curve (AUROC) and 95% confidence intervals (CI)^32^ for each biomarker, and CRP, to discriminate between patients with and without TB across both clinical syndromes, since we observed similar performance in each cohort. We compared AUROCs for each RNA signature to CRP using paired DeLong tests, with adjustment for multiple testing using a Benjamini-Hochberg correction. Sensitivity, specificity and predictive values for each biomarker were calculated at different test thresholds: two standard scores above the mean of the healthy control population (Z2) for the RNA signatures, 10 mg/L for CRP, and at the Youden Index of each ROC curve. In order to benchmark test performance against the WHO TPP^10^, we also calculated the specificity and predictive value of each biomarker at thresholds that achieved 90% sensitivity.

### Statistical power

The sample size of each cohort was primarily determined by the number of participants in the parent studies for which blood RNA, CRP and reference standard results were available. We calculated statistical precision in PASS 2024 (Power Analysis and Sample Size Software, National Council for Social Studies), using published models for estimates of sample size calculations in diagnostic tests^33,34^. Our sample sizes provided >80% power to test whether each biomarker could achieve the WHO TPP thresholds of 90% sensitivity with a 5% confidence interval and 70% specificity with 10% confidence interval (Supplementary Figure 5).

### Secondary analyses

In secondary analyses, we evaluated biomarker performance stratified by disease site. We also performed subgroup analyses by HIV status, and by HIV treatment status for HIV positive cases. The clinical utility of candidate TB screening strategies was assessed by decision curve analysis using the ‘rmda’ package as previously described^12,23^ to evaluate net benefit for the best performing blood RNA biomarker (at a threshold of Z2) to guide confirmatory testing, in comparison to confirmatory testing for all, confirmatory testing for none, and confirmatory testing guided by CRP (at thresholds of ≥5mg/L and ≥10mg/L). In addition to the alternative reference standard definitions, we also performed a sensitivity analysis without batch correction.

## Results

### Characteristics of the study cohorts

The pooled cohort comprised 440 individuals, of whom 66 people were excluded because they had any one of missing microbiological, CRP or RNA results (Supplementary Table 1). The excluded participants were generally comparable to those included in the analysis (Supplementary Table 1). Among the 374 participants included in the analysis, .275 had lymphadenitis and 99 had pericarditis (Table 1, Figure 1). Median age was 38 years (interquartile range 31-48), 181/374 (48%) were female and 200 (54%) were PLWH, of whom 95/198 (48%) with known antiretroviral therapy status were receiving antiretroviral therapy at enrolment.

**Figure 1.**
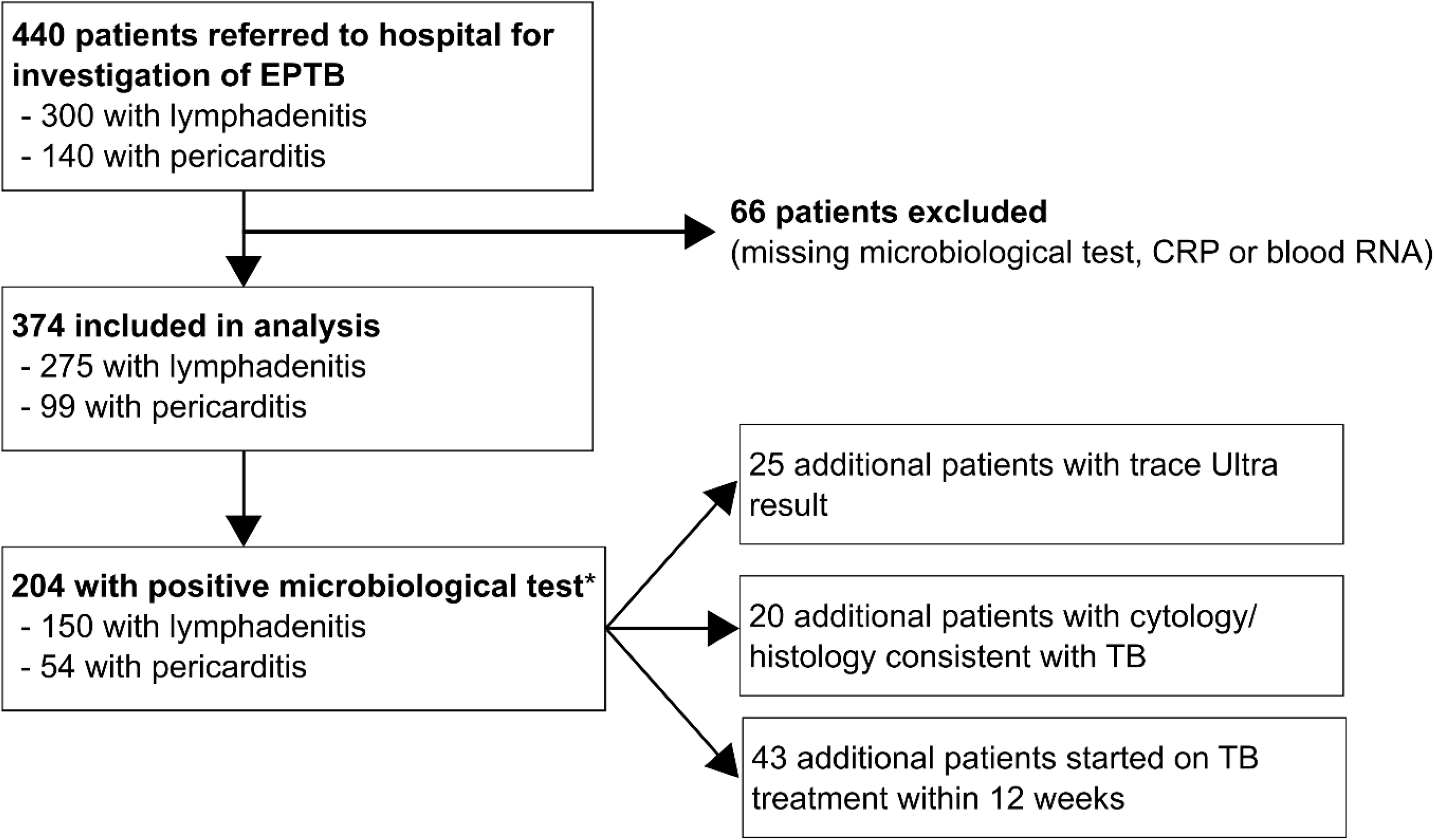
Study cohort consort diagrams. Consort diagrams for recruitment and analysis of study participants under investigation for lymphadenitis or pericarditis.

**Table 1.** Summary characteristics of lymphadenitis and pericarditis cohorts.

| Characteristic |  | Overall<br>N = 374 | Lymph node<br>N = 275 | Pericardial<br>N = 99 |
| --- | --- | --- | --- | --- |
| Age (years) |  | 38 (31, 48) | 38 (31, 46) | 37 (30, 49) |
| Gender | Female | 181 (48%) | 138 (50%) | 43 (43%) |
|  | Male | 193 (52%) | 137 (50%) | 56 (57%) |
| Previous TB | Yes | 91 (24%) | 70 (26%) | 21 (21%) |
|  | Missing | 2 | 2 | 0 |
| HIV status | Negative | 172 (46%) | 131 (48%) | 41 (41%) |
|  | Positive | 200 (54%) | 142 (52%) | 58 (59%) |
|  | Missing | 2 | 2 | 0 |
| On antiretroviral therapy* | Yes | 95 (48%) | 75 (54%) | 20 (34%) |
|  | Missing | 2 | 2 | 0 |
| CD4 (cells/mm<sup>3</sup>) | Yes | 158 (49, 288) | 158 (46, 272) | 176 (52, 332) |
|  | Missing | 175 | 133 | 42 |
| CD4 <200 (cells/mm<sup>3</sup>) | Yes | 118 (59%) | 84 (59%) | 34 (60%) |
|  | Missing | 175 | 133 | 42 |
| Site of disease molecular tests (N) | 0 | 2 (0.5%) | 2 (0.7%) | 0 (0%) |
|  | 1 | 156 (42%) | 156 (57%) | 0 (0%) |
|  | 2 | 124 (33%) | 117 (43%) | 7 (7.1%) |
|  | 3 | 64 (17%) | 0 (0%) | 64 (65%) |
|  | 4 | 28 (7.5%) | 0 (0%) | 28 (28%) |
| Fluid Xpert or Ultra | Negative | 154 (41%) | 117 (43%) | 37 (37%) |
|  | Positive | 218 (59%) | 156 (57%) | 62 (63%) |
|  | Not done | 2 | 2 | 0 |
| Fluid culture | Negative | 174 (67%) | 121 (74%) | 53 (56%) |
|  | Positive | 84 (33%) | 42 (26%) | 42 (44%) |
|  | Not done | 116 | 112 | 4 |
| Biopsy culture | Negative | 8 (38%) | 0 (NA%) | 8 (38%) |
|  | Positive | 13 (62%) | 0 (NA%) | 13 (62%) |
|  | Not done | 353 | 275 | 78 |
| Cytology consistent with TB | Yes | 126 (36%) | 126 (49%) | 0 (0%) |
|  | Not done | 28 | 16 | 12 |
| Histology consistent with TB | Yes | 9 (39%) | 0 (NA%) | 9 (39%) |
|  | Not done | 351 | 275 | 76 |
| Sputum culture or molecular | Negative | 39 (66%) | 22 (63%) | 17 (71%) |
|  | Positive | 20 (34%) | 13 (37%) | 7 (29%) |
|  | Not done | 315 | 240 | 75 |
| Culture or molecular test positive (site or sputum) |  | 204 (55%) | 150 (55%) | 54 (55%) |
| Started TB treatment | Yes | 221 (61%) | 143 (54%) | 78 (79%) |
|  | Missing | 9 | 9 | 0 |

204/374 (55%) of included participants met the primary reference standard for TB of a positive culture or molecular test for Mtb. Of these, 20/204 (10%) had positive Mtb sputum culture or sputum molecular tests. The prevalence of microbiologically confirmed TB was 150/275 (55%) among people with lymphadenitis, and 54/99 (55%) among those with pericarditis. In the lymphadenitis cohort, the majority of people with known site of disease (199/273; 73%) had cervical lymphadenitis, of whom 122/190 (61%) met the primary reference standard.

### Diagnostic accuracy of blood RNA signatures and CRP for pooled analysis of Tb lymphadenitis and pericarditis

In the primary analysis, all blood RNA biomarkers showed similar diagnostic accuracy to identify TB cases with microbiologically confirmed TB, with AUROC point estimates ranging 0.77-0.82 (Figure 2, Table 2). All the blood RNA biomarkers achieved statistically better AUROC than CRP with adjusted p-values of <0.05. The Roe3 signature achieved the highest AUROC point estimate of 0.82 (95% CI 0.77-0.86). By comparison, CRP achieved an AUROC of 0.61 (0.56-0.67). At pre-defined Z2 test thresholds for the blood RNA signatures, Roe3 achieved 78% (72-83) sensitivity and 69% (62-75) specificity. A pre-defined test threshold of 10 mg/L for CRP, which we have assessed previously as a triage test for pulmonary TB^12^, achieved a sensitivity of 88.3% (77-87) and specificity of 35% (29-43). When fixing the sensitivity of each test to 90% (in line with the WHO TPP), Roe3 achieved 52% (45-60) specificity and CRP achieved 27% (21-34) specificity, both below the WHO specificity target of 70% (Supplementary Table 2).

**Figure 2.**
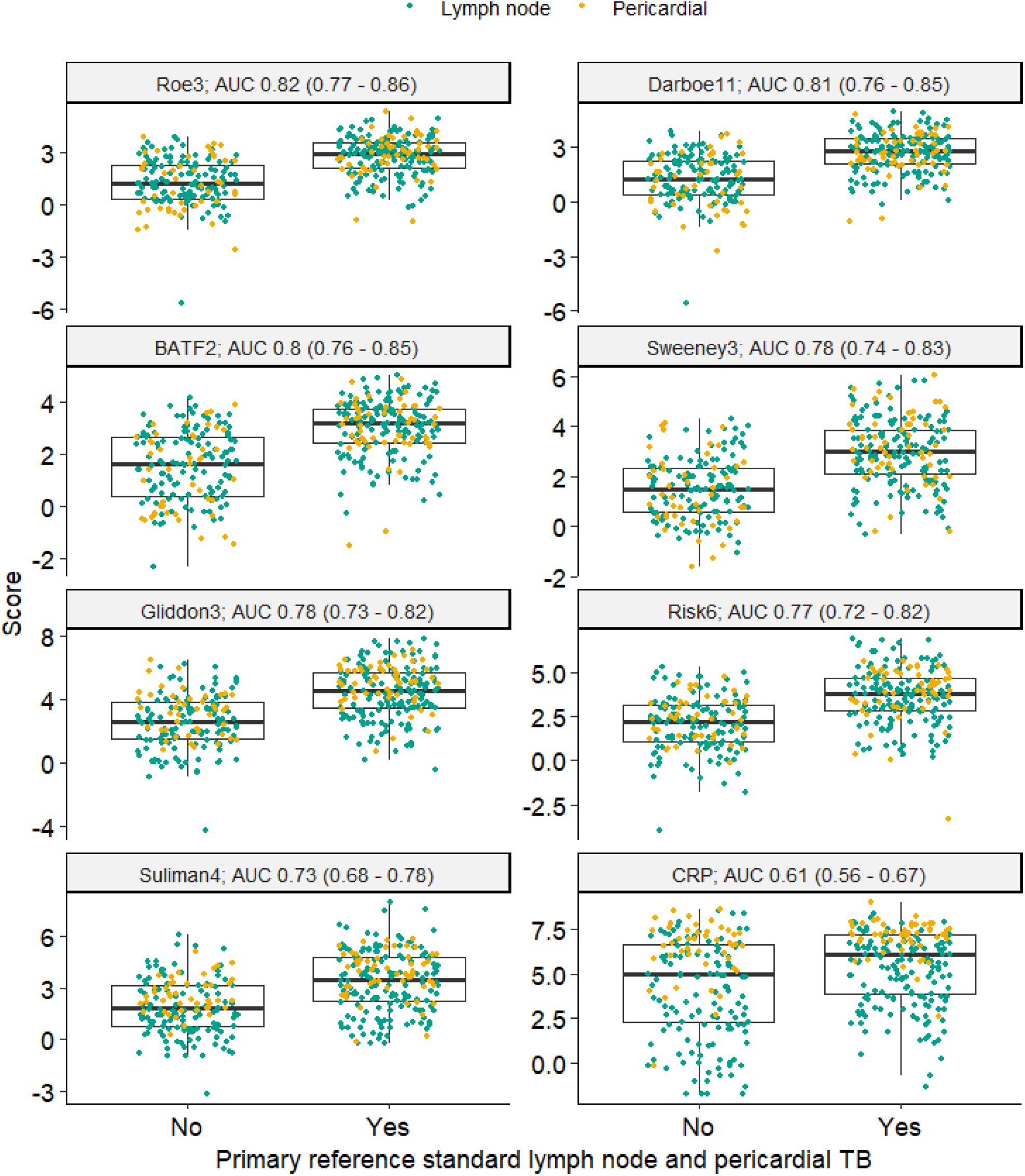
Blood RNA biomarker discrimination of primary reference standard for lymph node and pericardial TB. Scores shown as Z-scores for RNA signatures, and log-2 transformed CRP (mg/L), stratified by disease cohort (green = presumptive lymphadenitis and yellow = presumptive pericarditis). Discrimination of the pooled data set presented as area under the receiver operating characteristic curve (AUC), with 95% confidence intervals.

**Table 2.**
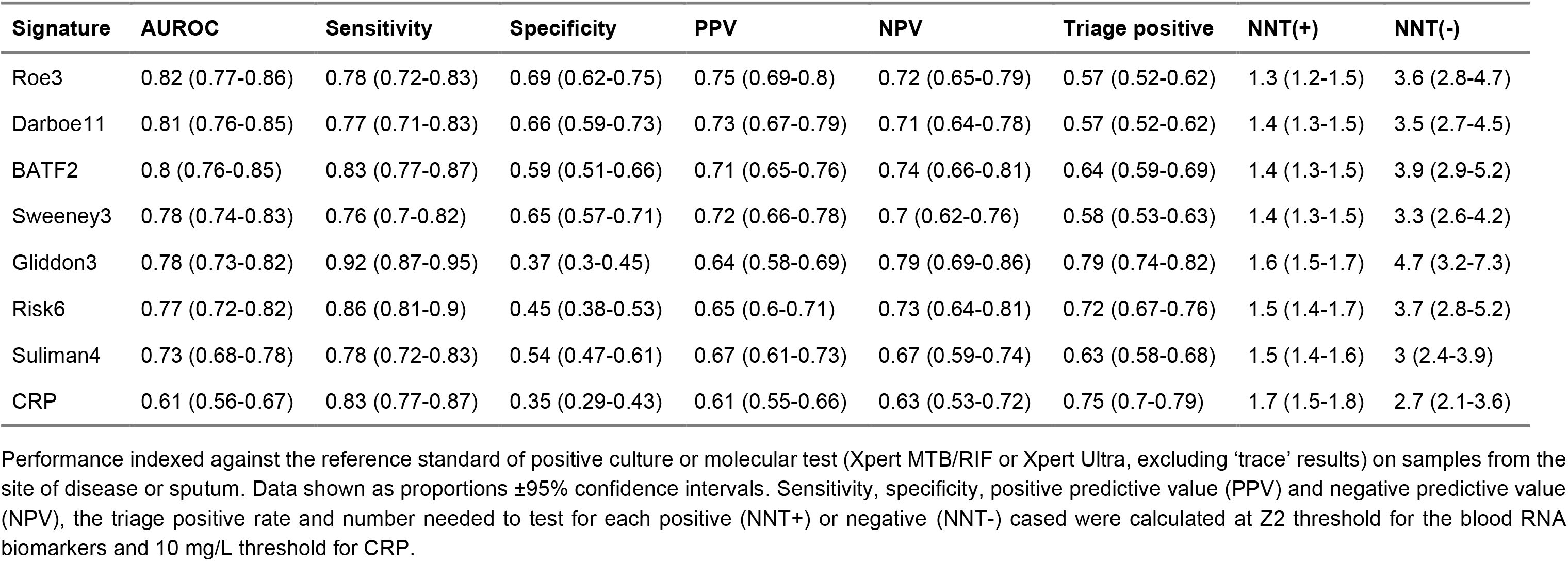
Diagnostic accuracy of blood RNA biomarkers and CRP for TB lymphadenitis and pericarditis.

| **Signature** | **AUROC** | **Sensitivity** | **Specificity** | **PPV** | **NPV** | **Triage positive** | **NNT(+)** | **NNT(-)** |
| --- | --- | --- | --- | --- | --- | --- | --- | --- |
| Roe3 | 0.82 (0.77-0.86) | 0.78 (0.72-0.83) | 0.69 (0.62-0.75) | 0.75 (0.69-0.8) | 0.72 (0.65-0.79) | 0.57 (0.52-0.62) | 1.3 (1.2-1.5) | 3.6 (2.8-4.7) |
| Darboe11 | 0.81 (0.76-0.85) | 0.77 (0.71-0.83) | 0.66 (0.59-0.73) | 0.73 (0.67-0.79) | 0.71 (0.64-0.78) | 0.57 (0.52-0.62) | 1.4 (1.3-1.5) | 3.5 (2.7-4.5) |
| BATF2 | 0.8 (0.76-0.85) | 0.83 (0.77-0.87) | 0.59 (0.51-0.66) | 0.71 (0.65-0.76) | 0.74 (0.66-0.81) | 0.64 (0.59-0.69) | 1.4 (1.3-1.5) | 3.9 (2.9-5.2) |
| Sweeney3 | 0.78 (0.74-0.83) | 0.76 (0.7-0.82) | 0.65 (0.57-0.71) | 0.72 (0.66-0.78) | 0.7 (0.62-0.76) | 0.58 (0.53-0.63) | 1.4 (1.3-1.5) | 3.3 (2.6-4.2) |
| Gliddon3 | 0.78 (0.73-0.82) | 0.92 (0.87-0.95) | 0.37 (0.3-0.45) | 0.64 (0.58-0.69) | 0.79 (0.69-0.86) | 0.79 (0.74-0.82) | 1.6 (1.5-1.7) | 4.7 (3.2-7.3) |
| Risk6 | 0.77 (0.72-0.82) | 0.86 (0.81-0.9) | 0.45 (0.38-0.53) | 0.65 (0.6-0.71) | 0.73 (0.64-0.81) | 0.72 (0.67-0.76) | 1.5 (1.4-1.7) | 3.7 (2.8-5.2) |
| Suliman4 | 0.73 (0.68-0.78) | 0.78 (0.72-0.83) | 0.54 (0.47-0.61) | 0.67 (0.61-0.73) | 0.67 (0.59-0.74) | 0.63 (0.58-0.68) | 1.5 (1.4-1.6) | 3 (2.4-3.9) |
| CRP | 0.61 (0.56-0.67) | 0.83 (0.77-0.87) | 0.35 (0.29-0.43) | 0.61 (0.55-0.66) | 0.63 (0.53-0.72) | 0.75 (0.7-0.79) | 1.7 (1.5-1.8) | 2.7 (2.1-3.6) |

Performance indexed against the reference standard of positive culture or molecular test (Xpert MTB/RIF or Xpert Ultra, excluding 'trace' results) on samples from the site of disease or sputum. Data shown as proportions ± 95% confidence intervals. Sensitivity, specificity, positive predictive value (PPV) and negative predictive value (NPV), the triage positive rate and number needed to test for each positive (NNT+) or negative (NNT-) cased were calculated at Z2 threshold for the blood RNA biomarkers and 10 mg/L threshold for CRP.

### Secondary analyses

In secondary analyses, we evaluated the performance of the biomarkers stratified by disease cohort. We found their performance to be comparable in lymphadenitis and pericarditis (Supplementary Figure 6-7, Supplementary Table 3-4). Of note, among TB cases, the CRP levels in the pericarditis cohort were significantly higher than that of lymphadenitis cases. This difference was less evident among the blood RNA signatures (Supplementary Figure 8). Biomarker diagnostic accuracy using alternate reference standards was similar to that of the primary analysis (Supplementary Figure 9-12). The point estimates for biomarker diagnostic accuracy among PLWH were lower than those without HIV infection but did not reach statistically significant differences (Supplementary Table 5).

In addition, we used the pooled dataset to evaluate clinical utility of Roe3 and that of CRP, using decision curve analysis, benchmarked against the alternative approaches of undertaking confirmatory diagnostic investigations in all or none^35^. This analysis quantifies the net benefit of a given approach, representing the trade-off between the true positive rate and a weighted false positive rate across a range of test threshold probabilities that trigger an intervention. The test threshold probabilities (reflecting the perceived cost:benefit ratio that triggers an intervention) are used for weighting the false positive rate. The intervention triggered by a triage test would be to proceed to confirmatory diagnostic investigations. Therefore, the triage test threshold probability can be considered as the number willing to investigate to identify each TB case. The Roe3 blood RNA biomarker (>Z2) achieved greater net benefit than confirmatory testing for all at a test threshold probability >0.28. By comparison CRP (>10 mg/L) provided greater net benefit than confirmatory testing for all at a test threshold probability >0.37 (Figure 3). These data suggest that using blood RNA biomarkers to direct use of confirmatory testing provides the best approach for comparable services in HIV/TB hyperendemic settings where the number willing to test by confirmatory diagnostics is fewer than 4 for each case of EPTB. In settings willing to test more than 4 people with a confirmatory test for each true case of EPTB, a test all approach achieves greater net benefit than a triage approach using any of the biomarkers tested in the present study.

**Figure 3.**
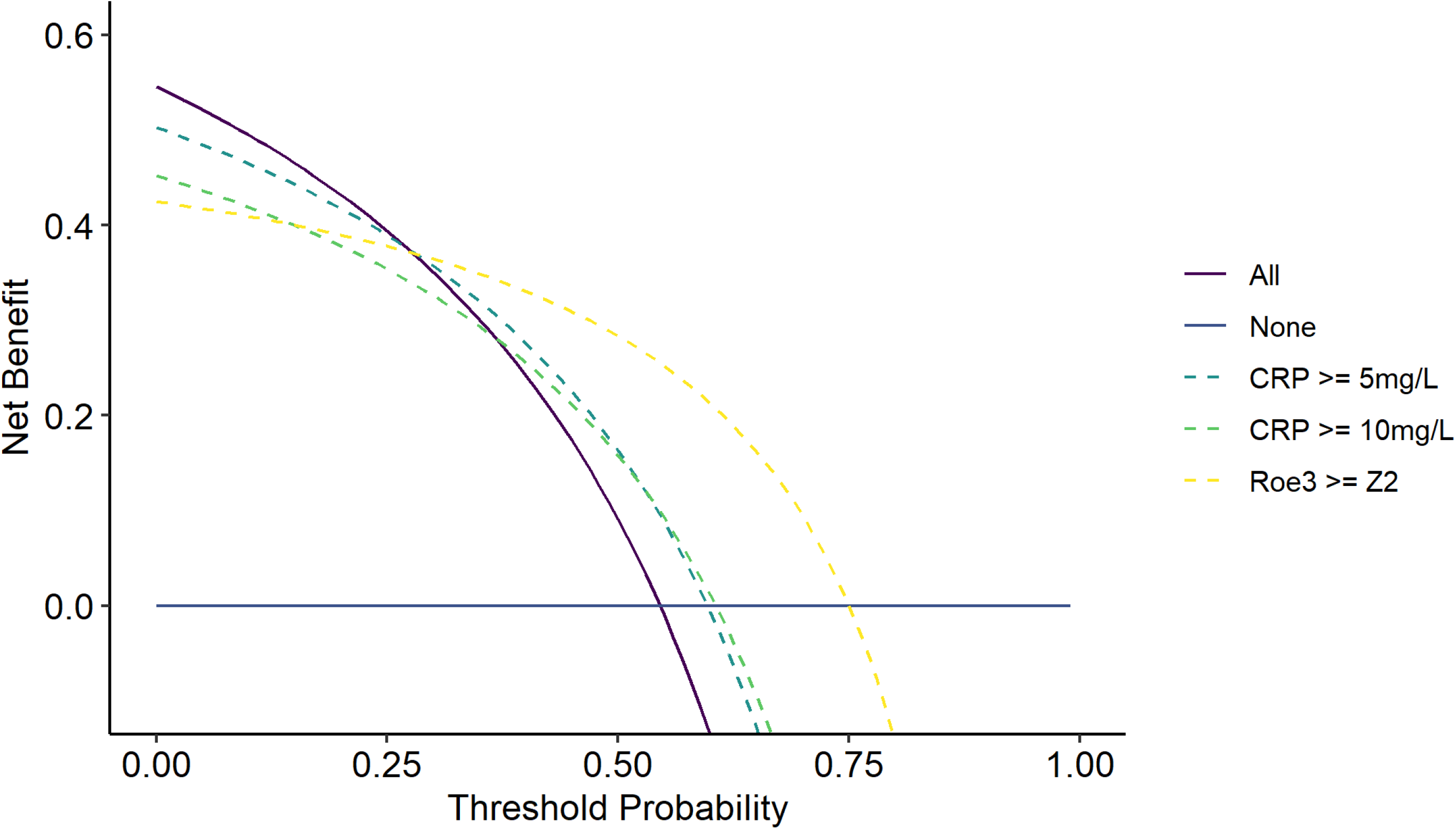
Decision curve analysis for alternative triage strategies to trigger confirmatory investigations for TB. Net benefit (true positive rate - false positive rate weighted by cost:benefit ratio) for investigate all and investigate none approaches across the range of threshold probabilities that a service may use to trigger confirmatory investigations for TB is compared to that of decisions to investigate triggered by each of triage strategies indicated: CRP (≥5 and ≥10mg/L), and Roe (Z2).

## Discussion

To the best of our knowledge, our findings provide the first diagnostic accuracy study of blood RNA biomarkers focusing on extrapulmonary TB. We undertook our study in a hyperendemic setting for both TB and HIV. In a pooled data set of people under investigation for TB lymphadenitis or pericarditis, all the blood RNA biomarkers tested had comparable performance with similar AUROCs as previously reported for pulmonary TB^11,36^. For most biomarkers, blood RNA signatures showed higher point estimates for accuracy in people without HIV compared to in PLWH, but with overlapping confidence intervals. In addition, we undertook head-to-head comparison of blood RNA biomarkers to serum CRP concentrations. In contrast to previous reports, where CRP measurements achieve similar diagnostic accuracy to blood RNA biomarkers in pulmonary TB^12,23^, we found CRP was significantly inferior to blood RNA biomarkers for the EPTB syndromes tested here, both among PLWH and people without HIV.

Importantly sputum tests identified less than 10% of the TB cases in our pooled analysis of TB lymphadenitis and pericarditis, often because sputum acquisition was not attempted. Given that people with extrapulmonary TB may have concomitant pulmonary TB, this highlights both the importance of strengthening sputum collection in addition to sampling from the site of disease in EPTB. Therefore, we envisioned the primary application of biomarkers of EPTB as triage tests to prioritise use of invasive sampling for confirmatory testing. The application of such tests in a primary care setting may prioritise patients for referral to secondary care reducing costs and attrition across the cascade of care. In this context, none of the biomarkers tested met the combined sensitivity of >90% and specificity >70% recommended by WHO^10^. Nonetheless, we assessed their potential clinical utility by decision curve analysis. The best performing blood RNA biomarker showed greater net benefit than either CRP or unrestricted approaches to trigger invasive confirmatory testing for services aiming to reduce the number needed to investigate to less than 4 to identify each case of TB. In settings with such high TB prevalence and therefore high prior probability of TB, unrestricted approaches to confirmatory testing may be preferable. Settings with lower prior probability of TB may be expected to derive greater benefit from triage tests.

In our secondary analyses, we showed that the performance of the biomarkers was equivalent in TB lymphadenitis and pericarditis. In addition, the use of alternative reference standards did not significantly alter our results. We focused on selected blood RNA biomarkers, previously identified by systematic review, which showed the best performance for diagnosis of pre-clinical/incipient as well as symptomatic TB and were sufficiently concise to undertake multiplex measurements using a validated Nanostring platform^16,23,36^. We have previously shown in a diagnostic accuracy study for pulmonary TB among PLWH that these RNA biomarkers show greater correlation to each other than to CRP^23^, suggesting that TB associated blood transcriptional changes are regulated differently to that of CRP levels. This interpretation is consistent with the findings in the present study showing that CRP does not identify TB cases in EPTB as well as blood RNA biomarkers. It is interesting that CRP levels were substantially higher in TB pericarditis than TB lymphadenitis cases. This difference was absent or less significant among the blood RNA signatures, suggesting a hierarchy for recruitment of immunological pathways with increasing inflammatory manifestations of disease. We propose that TNF and IFN signalling, which we previously identified as drivers of the blood RNA biomarkers of TB^16^ are more conserved across the spectrum of TB disease-sites, than the immunological pathways that regulate CRP, more strongly linked to IL-6 signalling^37^. If so, then despite recent interest in adopting point-of-care CRP as a triage test for TB^12,15,38^ we may expect that blood RNA biomarkers provide a more consistent performance for the range of disease manifestations beyond pulmonary TB. Interestingly, IL-6 signalling has also been associated with risk of TB^39^ and its immunopathogenesis^31^ consistent with the finding that CRP levels are higher in TB pericarditis that reflects a more severe variant of disease than TB lymphadenitis.

The strengths of our study include robust classification of TB based on highly specific microbiological tests in the primary analysis, complemented by sensitivity analyses using alternative reference standards; head-to-head comparison of multiple blood RNA signatures with CRP; inclusion of two distinct EPTB syndromes; and adequate sample sizes to evaluate the performance of biomarkers with a good level of precision. Our study also has important limitations. The single site design may limit the generalisability of our findings, particularly for settings with lower prevalence of TB and HIV. Future studies of EPTB in different settings will be required to address this limitation. Settings with lower prior probability of TB, for example earlier in the cascade of care or lower prevalence settings may reveal much greater differential clinical utility between biomarker triage and unrestricted approaches to confirmatory testing for EPTB. In addition, we cannot confidently generalise our findings across the full spectrum of EPTB diseases. The similarities in RNA biomarker performance between TB lymphadenitis and TB pericarditis is a valuable starting point, but future studies will require to test a wider range of EPTB syndromes.

Notwithstanding these limitations, our study substantially extends our insights into the real-world evaluation of the leading candidates for TB diagnostic triage tests by providing the largest available data for EPTB. Next generation blood RNA biomarkers show some clinical utility, but fall short of the desired accuracy targets. Importantly, blood CRP measurements show significantly inferior accuracy to blood RNA biomarkers in EPTB, and to that previously reported accuracy in pulmonary TB. We conclude that CRP based triage is unlikely to be useful in this context. Multivariable models incorporating clinical, epidemiological and laboratory measurements are urgently needed to overcome the limitations of individual biomarker approaches, alongside further innovation in discovery of novel diagnostic biomarkers.

## Footnotes

### Author contributions

Conceived and designed the study: GT, MN

Sample and clinical data collection: SM, BR, GN, HM, BD, HT, ZP, SN, DM

Laboratory analysis: TM, SM, AC, PK

Data analysis: TM, SM, BR, RKG, CJC, AC, PK, GT, MN

Manuscript preparation: TM, SM, RKG, BR, GT, MN with input from all authors

### Data availability statement

Processed Nanostring and CRP data with accompanying metadata will be provided as supplementary data at the time of peer-reviewed publication.

