## Supplementary Figures and Tables for "Blood RNA signatures outperform CRP triage of tuberculosis lymphadenitis and pericarditis"

### Supplementary data

**Supplementary Figure 1**

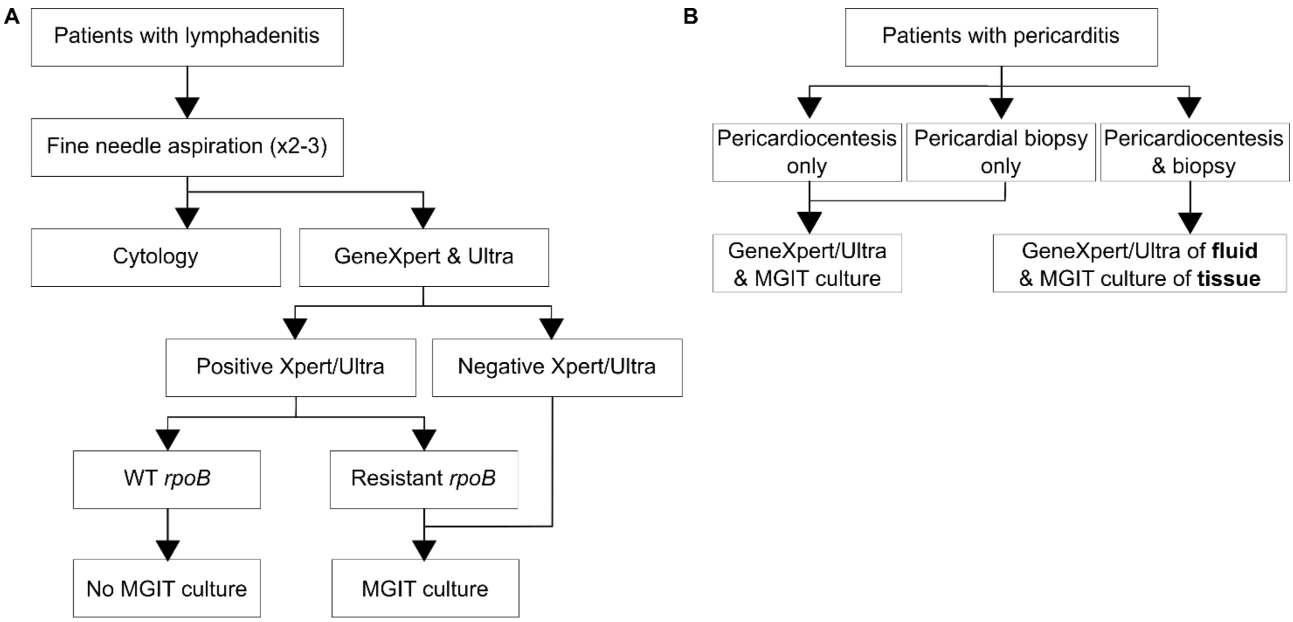

***Testing algorithm for patients with possible TB lymphadenitis or pericarditis***

**(A)** All patients under investigation for TB lymphadenitis had multipass fine needle aspiration. A sample for cytology was prioritised. The remaining sampling was used for PCR testing for Mtb using GeneXpert and Ultra. For those with negative PCR tests and those with positive PCR tests and rpoB mutations coferring rifampicin drug resistance any remaining sample was used for liquid culture using the Mycobacteria Growth Indicator Tube (MGIT960) platform. (B) All patients under investigation for TB pericarditis underwent sampling of pericardial fluid and or tissue biopsy. If only one sample type was collected, it was divided for both molecular testing and liquid culture. If both samples types were collected, the fluid was used for molecular testing and tissue biopsy was used for liquid culture.

**Supplementary Figure 2**

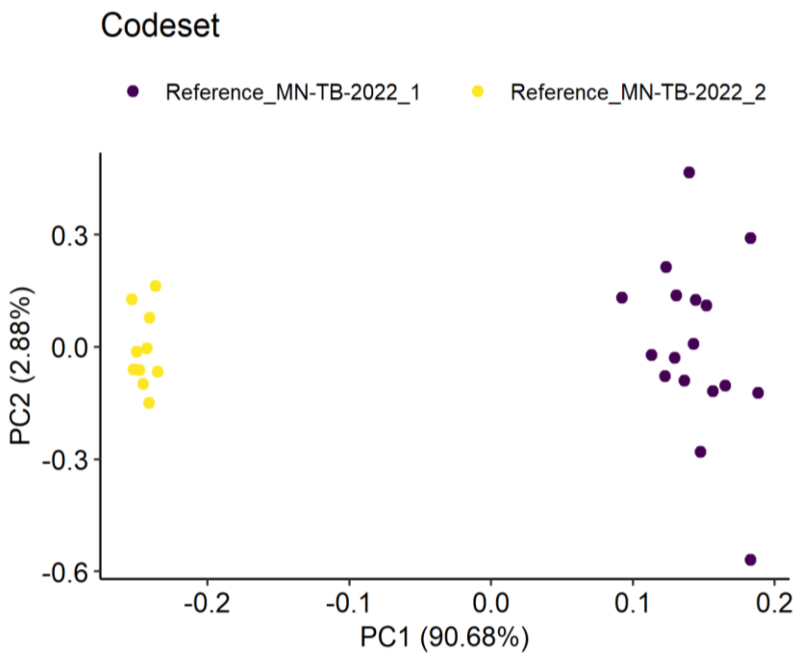

***Principal component analysis of Nanostring results from reference RNA sample.***

PCA clustering of standard reference RNA samples stratified by Nanostring codeset batch.

**Supplementary Figure 3**

***Distribution of gene expression data stratified by Nanostring codeset batch***

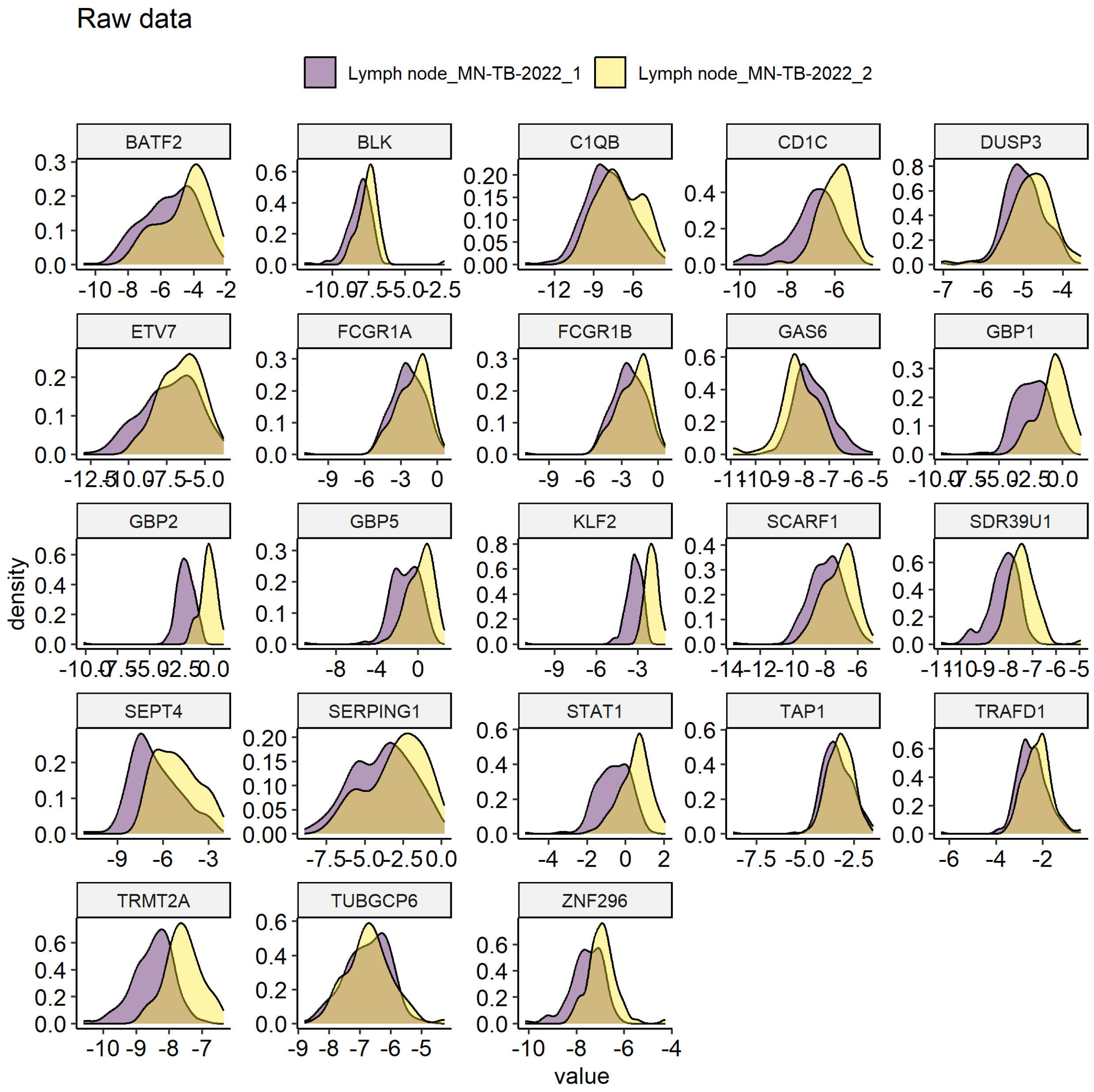

Gene level distributions of quantitative expression data in samples from lymphadenitis cohort stratified by Nanostring codeset batch.

**Supplementary Figure 4**

***Distributions of TB genes after ComBat***

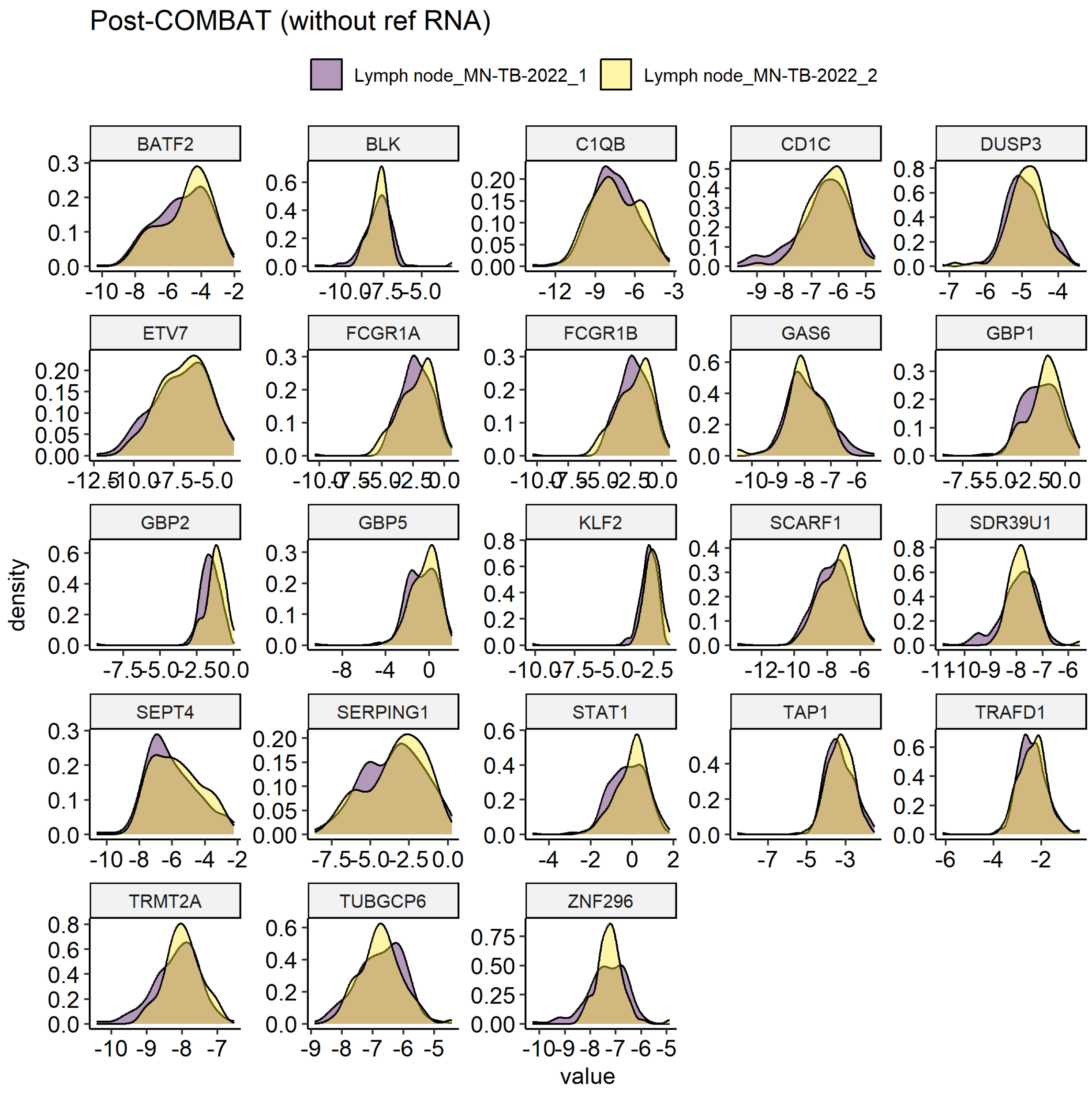

Gene level distributions of quantitative expression data in samples from lymphadenitis cohort stratified by Nanostring codeset batch following COMBAT batch correction.

**Supplementary Figure 5**

***Sample size calculations.***

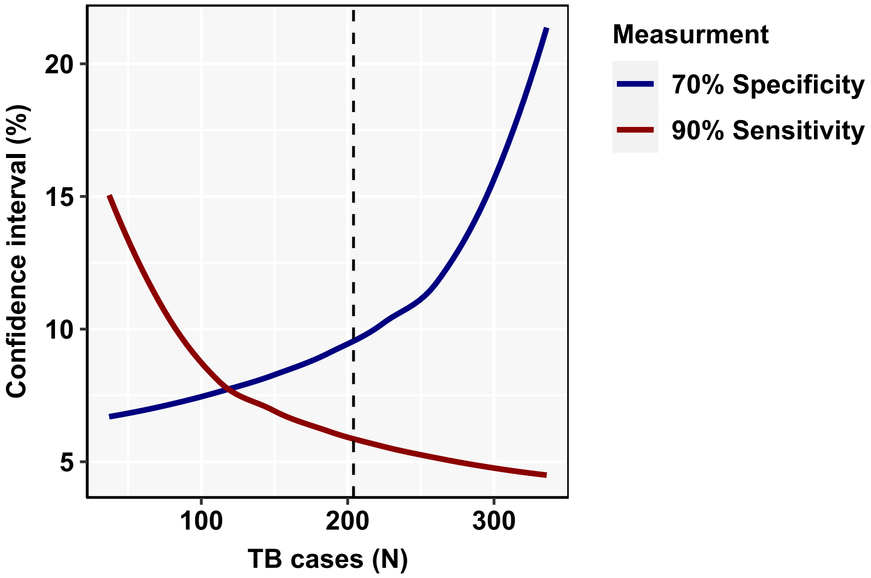

The relationship between confidence interval to detect a sensitivity of 90% and specificity of 70% is shown by number of TB cases for our total sample size (N=374) and power of >0.8, using PASS 2024 (Power Analysis and Sample Size Software, National Council for Social Studies). The dashed line shows the number of TB cases in our pooled data set (N=204).

**Supplementary Figure 6**

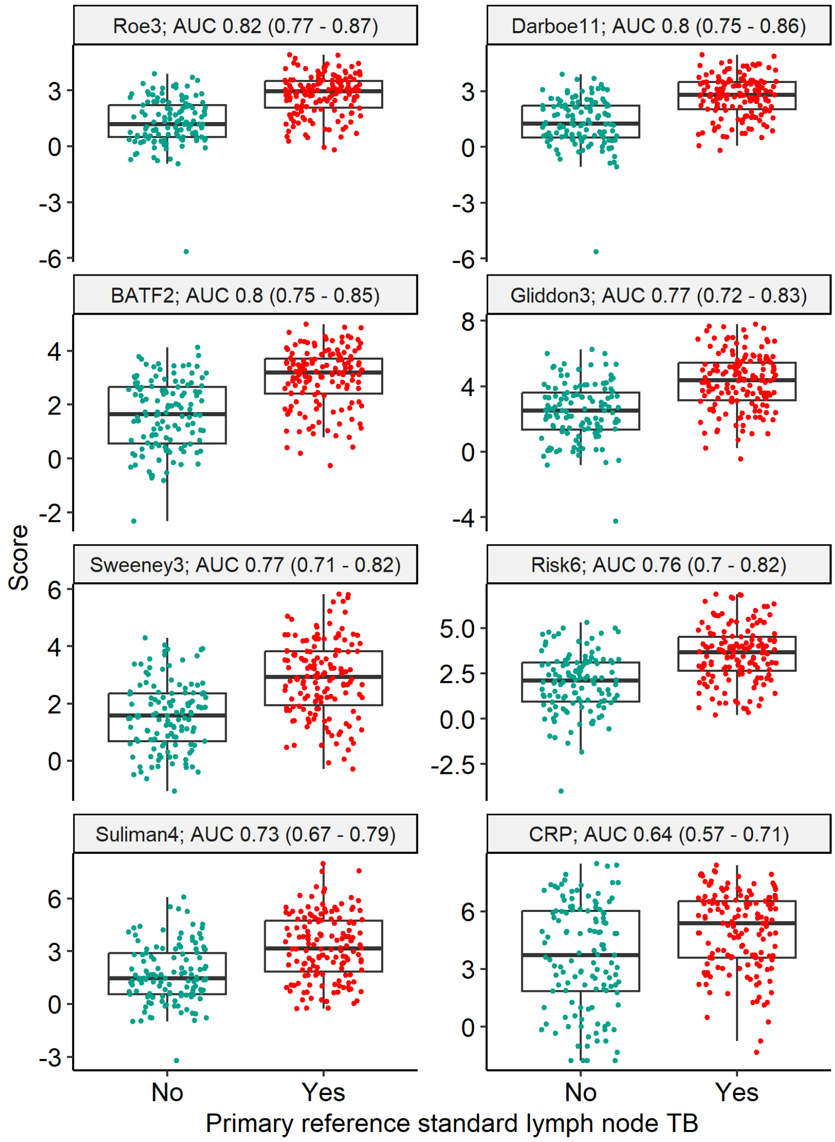

***Blood RNA biomarker discrimination of primary reference standard for lymph node TB.***

Scores are shown as Z-scores for RNA signatures, and log-2 transformed CRP (mg/L). Discrimination presented as area under the receiver operating characteristic curve (AUC), with 95% confidence intervals.

**Supplementary Figure 7**

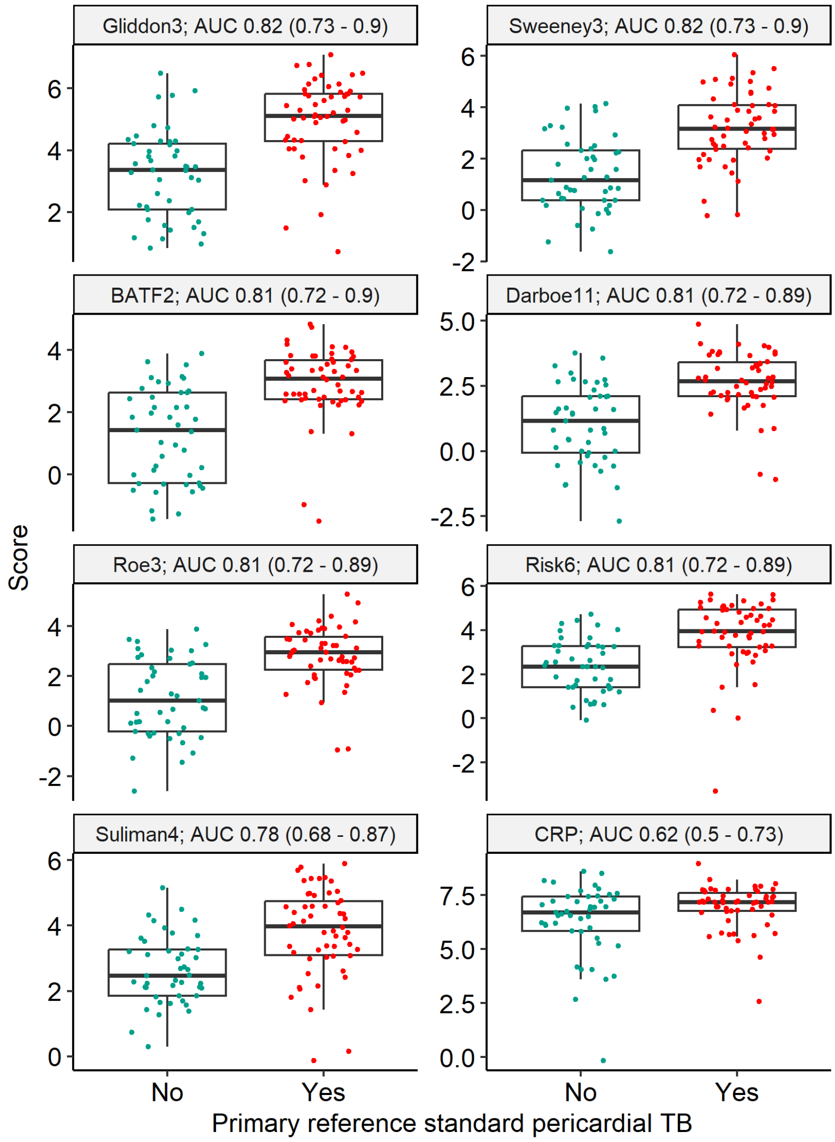

***Blood RNA biomarker discrimination of primary reference standard for pericardial TB.***

Scores are shown as Z-scores for RNA signatures, and log-2 transformed CRP (mg/L). Discrimination presented as area under the receiver operating characteristic curve (AUC), with 95% confidence intervals.

**Supplementary Figure 8**

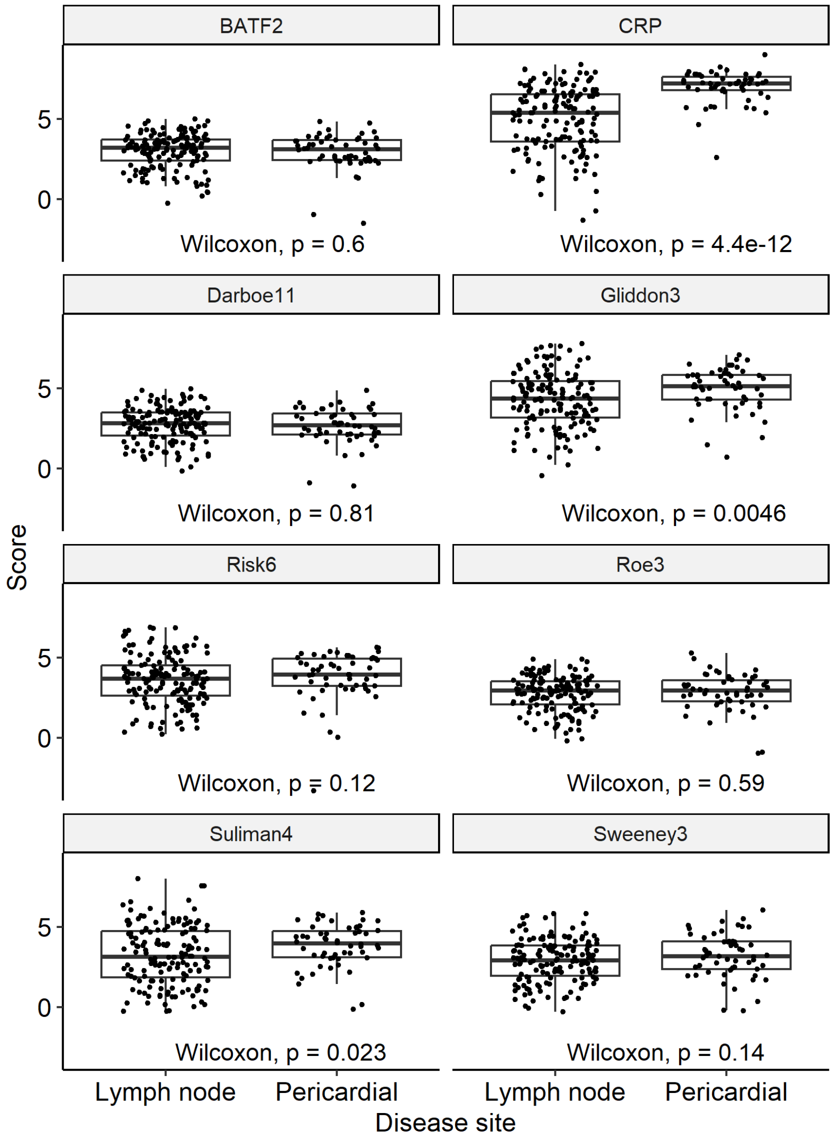

***Biomarker scores for TB lymphadenitis and pericarditis***

Z-scores for RNA signatures, and log-2 transformed CRP (mg/L) are shown for microbiologically confirmed TB stratified by disease site and compared by Wilcoxon Rank Test.

**Supplementary Figure 9**

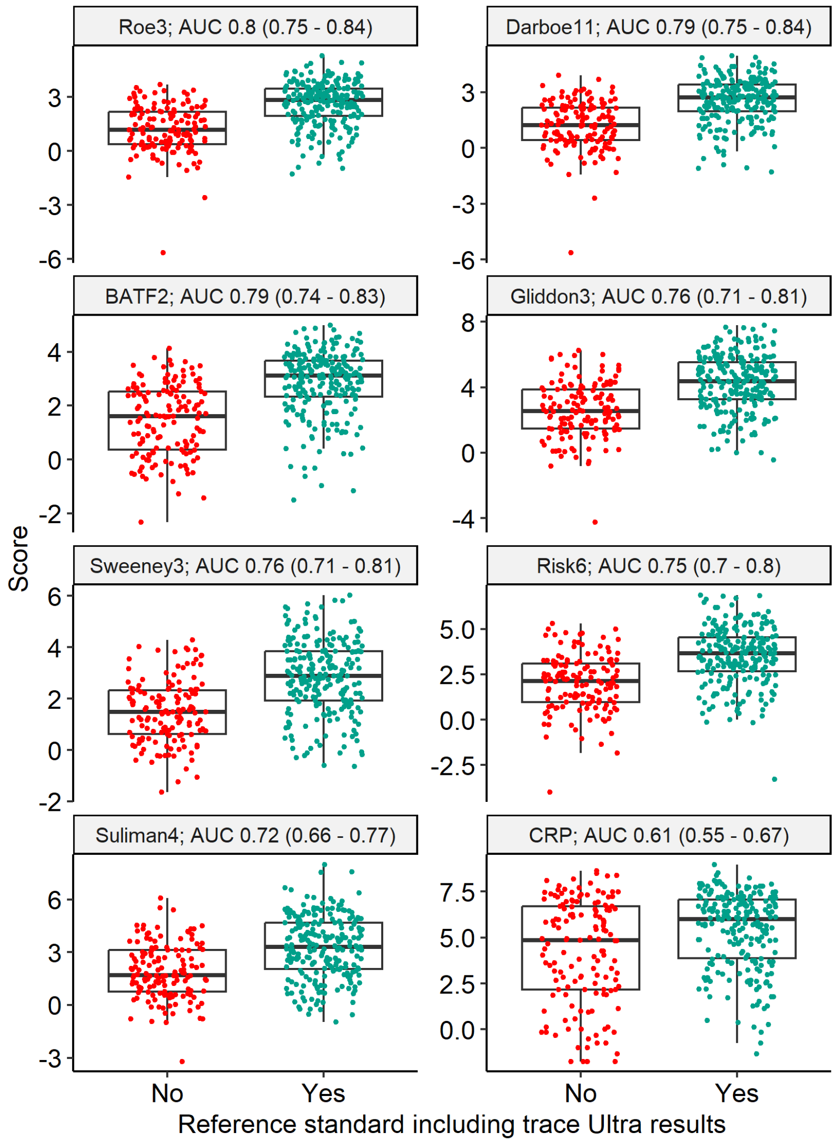

***Blood RNA biomarker discrimination of lymph node and pericardial TB including trace Ultra results.***

Scores are shown as Z-scores for RNA signatures, and log-2 transformed CRP (mg/L). Discrimination presented as area under the receiver operating characteristic curve (AUC), with 95% confidence intervals.

**Supplementary Figure 10**

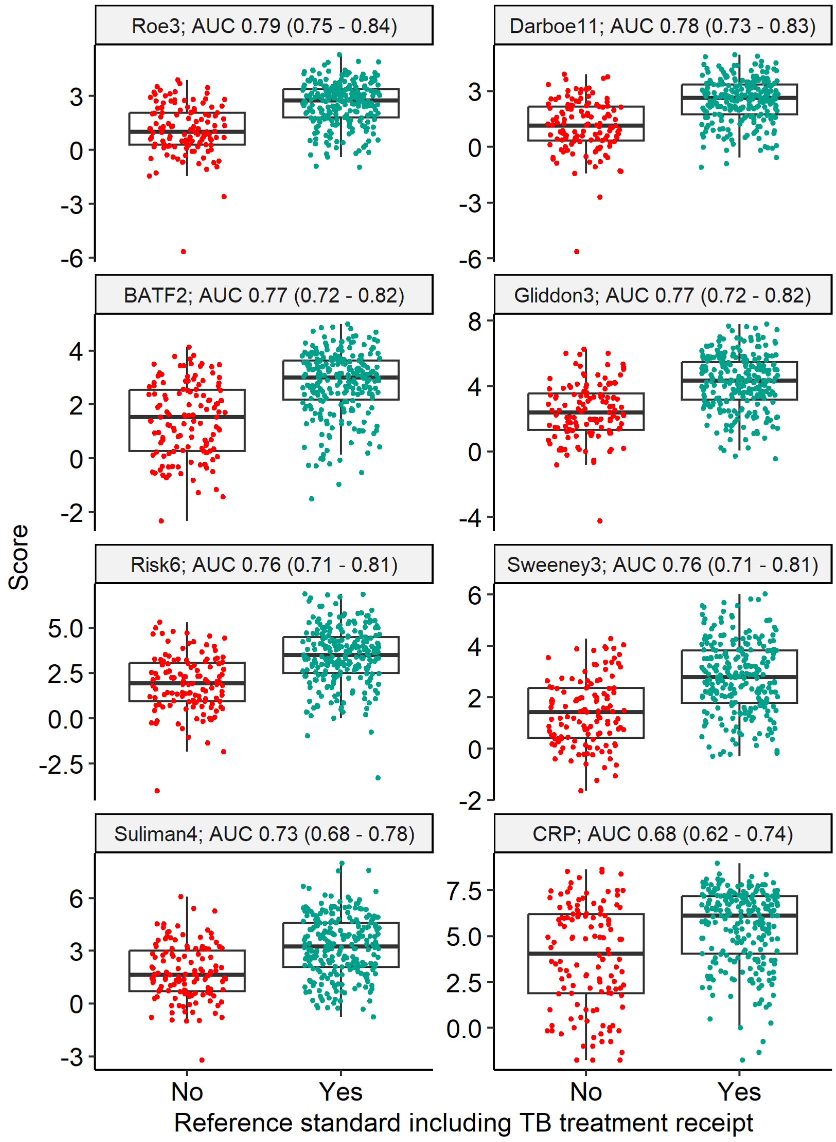

***Blood RNA biomarker discrimination of lymph node and pericardial TB including TB treatment initiation.***

Scores are shown as Z-scores for RNA signatures, and log-2 transformed CRP (mg/L). Discrimination presented as area under the receiver operating characteristic curve (AUC), with 95% confidence intervals.

**Supplementary Figure 11**

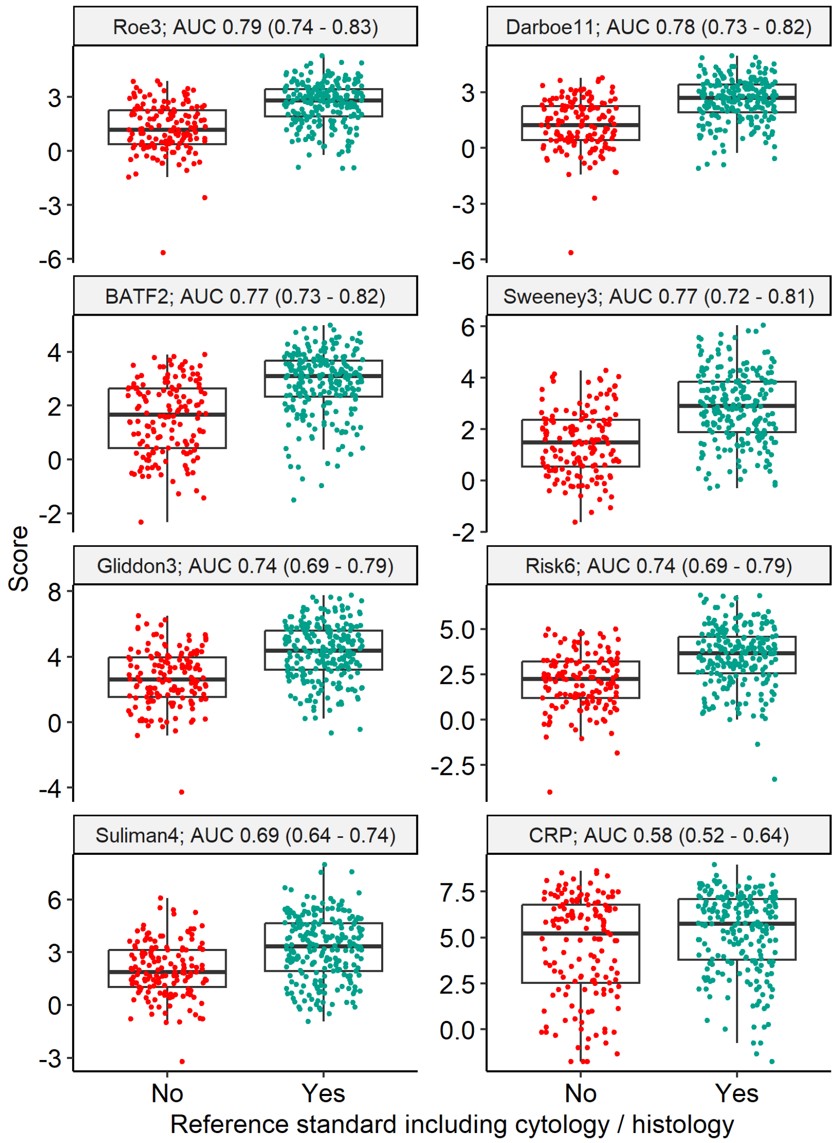

***Blood RNA biomarker discrimination of lymph node and pericardial TB including including cytology and histology diagnosis of TB.***

Scores are shown as Z-scores for RNA signatures, and log-2 transformed CRP (mg/L). Discrimination presented as area under the receiver operating characteristic curve (AUC), with 95% confidence intervals.

**Supplementary Figure 12**

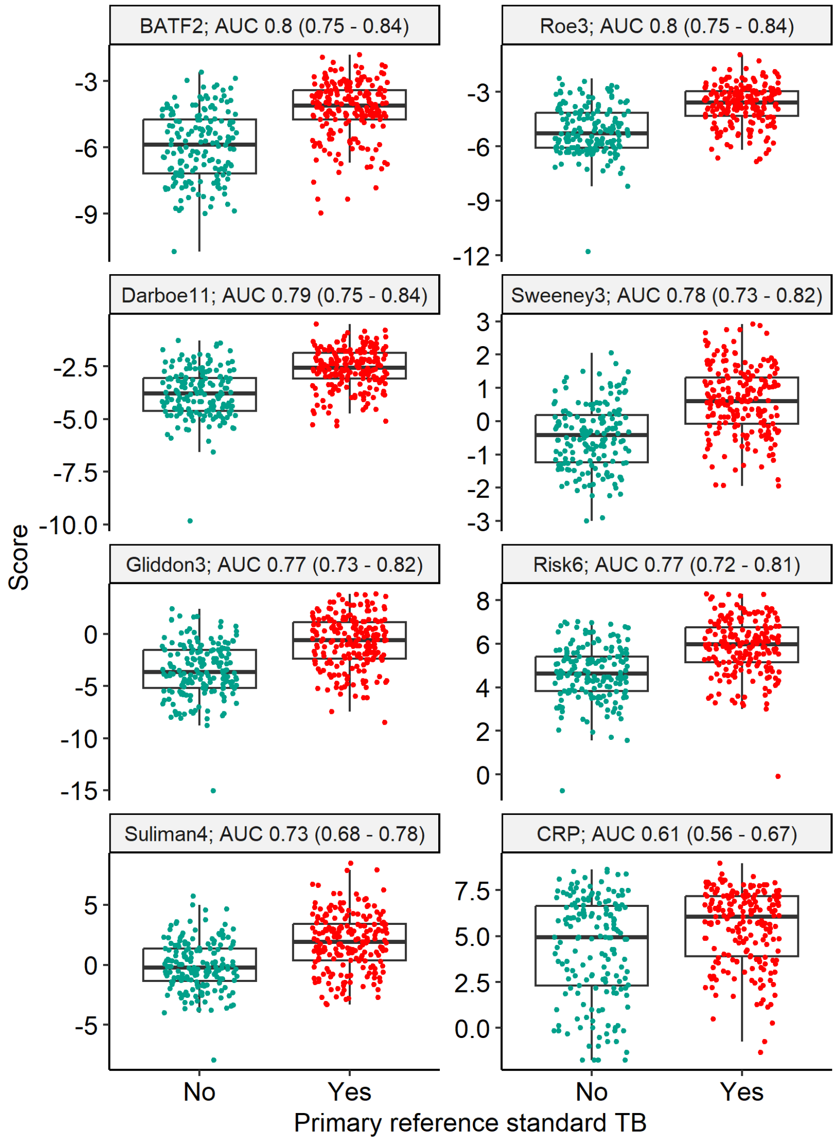

***Blood RNA biomarker discrimination of primary reference standard for lymph node and pericardial TB using raw Nanostring data without batch correction.***

Scores are shown as Z-scores for RNA signatures, and log-2 transformed CRP (mg/L). Discrimination presented as area under the receiver operating characteristic curve (AUC), with 95% confidence intervals.

**Supplementary Table 1**

***Baseline characteristics of the study cohort, stratified by inclusion***

| Characteristic | **Overall**, N = 440[1] | **Included**, N = 374[1] | **Excluded**, N = 66[1] |
|---|---|---|---|
| Age (years) | 38 (31, 48) | 38 (31, 48) | 39 (32, 49) |
| Gender | | | |
| Female | 218 (50%) | 181 (48%) | 37 (56%) |
| Male | 222 (50%) | 193 (52%) | 29 (44%) |
| Previous TB | 110 (25%) | 91 (24%) | 19 (30%) |
| Missing | 5 | 2 | 3 |
| HIV status | | | |
| Negative | 202 (46%) | 172 (46%) | 30 (47%) |
| Positive | 234 (54%) | 200 (54%) | 34 (53%) |
| Missing | 4 | 2 | 2 |
| CD4 (cells/mm3) | 157 (49, 287) | 158 (49, 288) | 154 (61, 275) |
| Missing | 207 | 175 | 32 |
| CD4 <200 cells/mm3 | 138 (59%) | 118 (59%) | 20 (59%) |
| Missing | 207 | 175 | 32 |
| On antiretroviral therapy | 107 (47%) | 95 (48%) | 12 (38%) |
| Missing | 210 | 176 | 34 |
| Site | | | |
| Lymph node | 300 (68%) | 275 (74%) | 25 (38%) |
| Pericardial | 140 (32%) | 99 (26%) | 41 (62%) |
| Culture or molecular positive (site or sputum) | 234 (53%) | 204 (55%) | 30 (47%) |
| Missing | 2 | 0 | 2 |
| Reason for exclusion | | | |
| No RNA data | 58 (88%) | 0 (NA%) | 58 (88%) |
| No CRP data | 6 (9.1%) | 0 (NA%) | 6 (9.1%) |
| No primary reference standard | 2 (3.0%) | 0 (NA%) | 2 (3.0%) |
| Missing | 374 | 374 | 0 |

[1]Median (IQR); n (%)

No systematic differences between included vs excluded participants. We included participants under investigation for presumptive lymph node or pericardial TB. Those with at least one molecular test result or TB culture from the site of disease, and CRP and RNA results available, were included.

**Supplementary Table 2**

***Blood RNA biomarker and CRP performance metrics for discrimination of primary reference standard in lymph node and pericardial TB, using alternative cut-offs.***

| Test | Cutoff | Sensitivity | Specificity | PPV | NPV | Triage positive | NNT(+) | NNT(-) |
|---|---|---|---|---|---|---|---|---|
| CRP | 5mg/L | 0.92 (0.88-0.95) | 0.25 (0.19-0.32) | 0.6 (0.54-0.65) | 0.73 (0.6-0.83) | 0.84 (0.8-0.88) | 1.7 (1.5-1.8) | 3.7 (2.5-5.7) |
| BATF2 | Maximal Youden Index | 0.72 (0.65-0.77) | 0.74 (0.66-0.8) | 0.76 (0.7-0.82) | 0.68 (0.61-0.75) | 0.51 (0.46-0.56) | 1.3 (1.2-1.4) | 3.2 (2.6-3.9) |
| CRP | Maximal Youden Index | 0.87 (0.82-0.91) | 0.33 (0.26-0.4) | 0.61 (0.55-0.66) | 0.68 (0.58-0.77) | 0.78 (0.74-0.82) | 1.6 (1.5-1.8) | 3.2 (2.4-4.4) |
| Gliddon3 | Maximal Youden Index | 0.6 (0.53-0.67) | 0.84 (0.78-0.89) | 0.82 (0.75-0.87) | 0.64 (0.57-0.7) | 0.4 (0.35-0.45) | 1.2 (1.1-1.3) | 2.8 (2.3-3.3) |
| Risk6 | Maximal Youden Index | 0.6 (0.53-0.66) | 0.84 (0.78-0.89) | 0.82 (0.75-0.87) | 0.64 (0.57-0.7) | 0.4 (0.35-0.45) | 1.2 (1.1-1.3) | 2.7 (2.3-3.3) |
| Roe3 | Maximal Youden Index | 0.71 (0.64-0.76) | 0.79 (0.72-0.84) | 0.8 (0.74-0.85) | 0.69 (0.62-0.75) | 0.48 (0.43-0.53) | 1.2 (1.2-1.4) | 3.2 (2.6-4) |
| Suliman4 | Maximal Youden Index | 0.75 (0.68-0.8) | 0.62 (0.54-0.69) | 0.7 (0.64-0.76) | 0.67 (0.59-0.74) | 0.58 (0.53-0.63) | 1.4 (1.3-1.6) | 3 (2.5-3.8) |
| Sweeney3 | Maximal Youden Index | 0.67 (0.6-0.73) | 0.79 (0.72-0.84) | 0.79 (0.73-0.85) | 0.67 (0.6-0.73) | 0.46 (0.41-0.51) | 1.3 (1.2-1.4) | 3 (2.5-3.7) |
| Darboe11 | Maximal Youden Index | 0.8 (0.74-0.85) | 0.65 (0.58-0.72) | 0.74 (0.67-0.79) | 0.74 (0.66-0.8) | 0.6 (0.55-0.64) | 1.4 (1.3-1.5) | 3.8 (2.9-5) |
| BATF2 | Sensitivity 90% | 0.9 (0.85-0.93) | 0.44 (0.37-0.51) | 0.66 (0.6-0.71) | 0.78 (0.69-0.86) | 0.75 (0.7-0.79) | 1.5 (1.4-1.7) | 4.6 (3.2-6.9) |
| CRP | Sensitivity 90% | 0.9 (0.85-0.93) | 0.27 (0.21-0.34) | 0.6 (0.54-0.65) | 0.69 (0.57-0.79) | 0.82 (0.78-0.86) | 1.7 (1.5-1.8) | 3.3 (2.4-4.8) |
| Gliddon3 | Sensitivity 90% | 0.9 (0.85-0.93) | 0.39 (0.32-0.47) | 0.64 (0.58-0.69) | 0.77 (0.67-0.84) | 0.77 (0.72-0.81) | 1.6 (1.4-1.7) | 4.3 (3-6.4) |
| Risk6 | Sensitivity 90% | 0.9 (0.85-0.93) | 0.38 (0.31-0.45) | 0.63 (0.58-0.69) | 0.76 (0.66-0.84) | 0.77 (0.73-0.81) | 1.6 (1.5-1.7) | 4.1 (2.9-6.1) |
| Roe3 | Sensitivity 90% | 0.9 (0.85-0.93) | 0.52 (0.45-0.6) | 0.69 (0.64-0.75) | 0.81 (0.73-0.88) | 0.71 (0.66-0.75) | 1.4 (1.3-1.6) | 5.4 (3.7-8) |
| Suliman4 | Sensitivity 90% | 0.9 (0.85-0.93) | 0.27 (0.21-0.34) | 0.6 (0.54-0.65) | 0.69 (0.57-0.79) | 0.82 (0.78-0.86) | 1.7 (1.5-1.8) | 3.3 (2.3-4.8) |
| Sweeney3 | Sensitivity 90% | 0.9 (0.85-0.93) | 0.42 (0.35-0.5) | 0.65 (0.59-0.71) | 0.78 (0.68-0.85) | 0.75 (0.71-0.79) | 1.5 (1.4-1.7) | 4.5 (3.2-6.7) |
| Darboe11 | Sensitivity 90% | 0.9 (0.85-0.93) | 0.45 (0.37-0.52) | 0.66 (0.6-0.71) | 0.79 (0.7-0.86) | 0.74 (0.7-0.78) | 1.5 (1.4-1.7) | 4.7 (3.3-7) |

**Supplementary Table 3**

***Blood RNA biomarker and CRP performance metrics for discrimination of primary reference standard in lymph node TB.***

| Signature | AUROC | Sensitivity | Specificity | PPV | NPV | Triage positive | NNT(+) | NNT(-) | p |
|---|---|---|---|---|---|---|---|---|---|
| Roe3 | 0.82 (0.77-0.87) | 0.76 (0.69-0.82) | 0.7 (0.61-0.77) | 0.75 (0.68-0.81) | 0.71 (0.62-0.78) | 0.55 (0.49-0.61) | 1.3 (1.2-1.5) | 3.4 (2.6-4.6) | <0.0001 |
| Darboe11 | 0.8 (0.75-0.86) | 0.75 (0.68-0.82) | 0.66 (0.58-0.74) | 0.73 (0.65-0.79) | 0.69 (0.6-0.77) | 0.56 (0.5-0.62) | 1.4 (1.3-1.5) | 3.2 (2.5-4.3) | <0.0001 |
| BATF2 | 0.8 (0.75-0.85) | 0.79 (0.72-0.85) | 0.58 (0.49-0.66) | 0.69 (0.62-0.76) | 0.7 (0.6-0.78) | 0.63 (0.57-0.68) | 1.4 (1.3-1.6) | 3.3 (2.5-4.5) | <0.0001 |
| Gliddon3 | 0.77 (0.72-0.83) | 0.91 (0.85-0.94) | 0.42 (0.33-0.5) | 0.65 (0.58-0.71) | 0.79 (0.67-0.87) | 0.76 (0.71-0.81) | 1.5 (1.4-1.7) | 4.7 (3.1-7.6) | <0.0001 |
| Sweeney3 | 0.77 (0.71-0.82) | 0.75 (0.67-0.81) | 0.65 (0.56-0.73) | 0.72 (0.64-0.78) | 0.68 (0.59-0.76) | 0.57 (0.51-0.62) | 1.4 (1.3-1.6) | 3.1 (2.5-4.1) | <0.0001 |
| Risk6 | 0.76 (0.7-0.82) | 0.85 (0.78-0.9) | 0.46 (0.38-0.55) | 0.65 (0.59-0.72) | 0.72 (0.61-0.8) | 0.71 (0.65-0.76) | 1.5 (1.4-1.7) | 3.5 (2.6-5.1) | <0.0001 |
| Suliman4 | 0.73 (0.67-0.79) | 0.73 (0.65-0.79) | 0.64 (0.55-0.72) | 0.71 (0.63-0.77) | 0.66 (0.57-0.74) | 0.56 (0.5-0.62) | 1.4 (1.3-1.6) | 3 (2.3-3.8) | 0.002 |
| CRP | 0.64 (0.57-0.71) | 0.77 (0.7-0.83) | 0.46 (0.38-0.55) | 0.63 (0.56-0.7) | 0.63 (0.53-0.72) | 0.67 (0.61-0.72) | 1.6 (1.4-1.8) | 2.7 (2.1-3.6) | |

P values indicate pairwise comparisons to CRP, with multiple testing correction.

**Supplementary Table 4**

***Blood RNA biomarker and CRP performance metrics for discrimination of primary reference standard in pericardial TB.***

| Signature | AUROC | Sensitivity | Specificity | PPV | NPV | Triage positive | NNT(+) | NNT(-) | p |
|---|---|---|---|---|---|---|---|---|---|
| Gliddon3 | 0.82 (0.73-0.9) | 0.94 (0.85-0.98) | 0.24 (0.14-0.39) | 0.6 (0.49-0.7) | 0.79 (0.52-0.92) | 0.86 (0.78-0.91) | 1.7 (1.4-2) | 4.7 (2.1-13.2) | 0.012 |
| Sweeney3 | 0.82 (0.73-0.9) | 0.81 (0.69-0.9) | 0.64 (0.5-0.77) | 0.73 (0.61-0.83) | 0.74 (0.59-0.85) | 0.61 (0.51-0.7) | 1.4 (1.2-1.6) | 3.9 (2.4-6.9) | 0.012 |
| BATF2 | 0.81 (0.72-0.9) | 0.93 (0.82-0.97) | 0.62 (0.48-0.75) | 0.75 (0.63-0.84) | 0.88 (0.72-0.95) | 0.68 (0.58-0.76) | 1.3 (1.2-1.6) | 8 (3.6-20.1) | 0.016 |
| Risk6 | 0.81 (0.72-0.89) | 0.91 (0.8-0.96) | 0.42 (0.29-0.57) | 0.65 (0.54-0.75) | 0.79 (0.6-0.91) | 0.76 (0.66-0.83) | 1.5 (1.3-1.8) | 4.8 (2.5-10.8) | 0.016 |
| Roe3 | 0.81 (0.72-0.89) | 0.83 (0.71-0.91) | 0.67 (0.52-0.79) | 0.75 (0.63-0.84) | 0.77 (0.62-0.87) | 0.61 (0.51-0.7) | 1.3 (1.2-1.6) | 4.3 (2.6-7.9) | 0.016 |
| Darboe11 | 0.81 (0.72-0.89) | 0.83 (0.71-0.91) | 0.67 (0.52-0.79) | 0.75 (0.63-0.84) | 0.77 (0.62-0.87) | 0.61 (0.51-0.7) | 1.3 (1.2-1.6) | 4.3 (2.6-7.9) | 0.016 |
| Suliman4 | 0.78 (0.68-0.87) | 0.93 (0.82-0.97) | 0.27 (0.16-0.41) | 0.6 (0.49-0.7) | 0.75 (0.51-0.9) | 0.84 (0.75-0.9) | 1.7 (1.4-2) | 4 (2-9.8) | 0.016 |
| CRP | 0.62 (0.5-0.73) | 0.98 (0.9-1) | 0.04 (0.01-0.15) | 0.55 (0.45-0.65) | 0.67 (0.21-0.98) | 0.97 (0.91-0.99) | 1.8 (1.5-2.2) | 3 (1.3-58.5) | |

P values indicate pairwise comparisons to CRP, with multiple testing correction.

**Supplementary Table 5**

***Blood RNA biomarker and CRP performance metrics for discrimination of TB status, stratified by selected subgroups.***

| Signature | HIV-positive | HIV-negative | p |
|---|---|---|---|
| BATF2 | 0.8 (0.74-0.86) | 0.79 (0.73-0.86) | 0.954 |
| CRP | 0.62 (0.54-0.71) | 0.57 (0.48-0.65) | 0.846 |
| Gliddon3 | 0.78 (0.72-0.84) | 0.75 (0.68-0.83) | 0.926 |
| Risk6 | 0.78 (0.72-0.84) | 0.73 (0.66-0.81) | 0.846 |
| Roe3 | 0.81 (0.75-0.87) | 0.8 (0.74-0.87) | 0.954 |
| Suliman4 | 0.77 (0.7-0.83) | 0.65 (0.57-0.73) | 0.238 |
| Sweeney3 | 0.79 (0.73-0.86) | 0.75 (0.68-0.83) | 0.846 |
| Darboe11 | 0.8 (0.74-0.86) | 0.8 (0.73-0.87) | 0.954 |
| Signature | On ART | No ART | p |
| BATF2 | 0.74 (0.64-0.84) | 0.84 (0.76-0.91) | 0.185 |
| CRP | 0.56 (0.44-0.68) | 0.64 (0.51-0.77) | 0.381 |
| Gliddon3 | 0.69 (0.58-0.8) | 0.84 (0.76-0.92) | 0.084 |
| Risk6 | 0.69 (0.59-0.8) | 0.84 (0.76-0.92) | 0.084 |
| Roe3 | 0.77 (0.67-0.86) | 0.84 (0.77-0.92) | 0.24 |
| Suliman4 | 0.67 (0.56-0.78) | 0.85 (0.77-0.92) | 0.084 |
| Sweeney3 | 0.72 (0.62-0.82) | 0.85 (0.78-0.93) | 0.084 |
| Darboe11 | 0.74 (0.64-0.84) | 0.84 (0.76-0.91) | 0.185 |

Accuracy shown for primary reference standard as area under the receiver operating characteristic curve with 95% confidence intervals. P values indicate comparisons between strata for each signature.
